# Assessing COVID-19 Risk Factors in Toronto Using a Localized Spatio-Temporal Conditional Autoregressive Model

**DOI:** 10.64898/2026.02.03.26345488

**Authors:** Afia Amoako, Mabel Carabali, Erjia Ge, Ashleigh R Tuite, David N Fisman

**Author notes:** Corresponding Author Afia Amoako, MScPH. Coauthors Email Address.

## Abstract

**Purpose:** Most spatio-temporal models identify COVID-19 sociodemographic and socioeconomic risk factors using methods that assume a single spatial dependency pattern across the city, which may not reflect reality. The purpose of this study is to apply a spatially and temporally localized Bayesian model to identify COVID-19 risk factors that account for localized context.

**Methods:** For this study, a spatio-temporal localized Bayesian Hierarchical Model (ST-LCAR) was used to assess the relationships between population factors (age, sex, income, visible minority status, and education) and COVID-19 relative risk. The ST-LCAR model accounts for spatial and temporal autocorrelation through spatio-temporal random effects along with piecewise intercepts to capture step changes in relative risk patterns that might be reflective of underlying local contexts. This study focuses on the first four complete waves of the COVID-19 pandemic across Forward Sortation Areas (FSAs) in the City of Toronto.

**Results:** A 10-percentage-point increase in the proportion of residents who identify as visible minorities was associated with a 3% increase in COVID-19 relative risk; however, this association varied across different social contexts. On the other hand, a 10-percentage-point increase in the proportion of residents with post-secondary education was associated with a 22% decrease in relative risk. Beyond quantitative relationships, our model identified 3 times higher COVID-19 relative risk in the northwestern portion of the city, with patterns varying over time.

**Conclusion:** The different COVID-19 patterns in the city of Toronto may have been shaped by the complex and diverse social contexts, products of ingrained systems of structural inequities that influence the living, working, and economic conditions of city residents. Public health interventions and pandemic preparedness should integrate an equity-focused lens that considers the diverse social contexts across the city and how it shapes health outcomes.

## INTRODUCTION

The COVID-19 pandemic has been a dynamic experience for the City of Toronto’s diverse population. Beyond biological risk factors, increased risk of COVID-19 infection and adverse outcomes in Toronto have been linked to the residents’ sociodemographic and socioeconomic factors such as education levels, race, immigrant and income status, as well as housing environment. ^1–6^

Long-standing systemic inequities (structural racism, economic deprivation, stigma) have shaped the urban landscape of Toronto through changing national immigration and economic policies, urban planning policies, gentrification and housing development, resulting in pockets(neighbourhoods) of the city with differing social contexts, sometimes changing abruptly between adjacent areas ^1–3,7,8^ Social context—the social, economic, environmental, and cultural conditions of a place — are complex and dictate the housing, neighbourhood, and work environments of neighbourhoods, as well as the cultural makeup, social networks, services, and resources associated with exposure and vulnerability to infectious diseases.^9,10^

As a result, COVID-19 case patterns might have reflected these differential neighbourhood contexts. In our previous descriptive study, COVID-19 case hotspots were localized in parts of the city with more socially and economically marginalized residents, sometimes in close proximity to cold spots, which were localized in areas where more affluent and highly educated residents reside.^11^

Simultaneously, the unequal experience of COVID-19 in the city has been far from static, with clustering patterns of COVID-19 varying over space and time, owing to evolving public health interventions, more transmissible viral variants, and the introduction of vaccines. ^12^ Furthermore, this dynamism is not independent of social context, as social context might influence compliance with public health interventions, access to vaccination, and confidence in vaccination benefits. ^13,14^ Understanding COVID-19 risk, therefore, requires one to account for both the city’s localized social landscape and the pandemic temporal dynamics.

Bayesian hierarchical models provide a suitable framework for this purpose. They are multilevel models that can assess exposure-response relationships through covariates, uncertainty and spatial structure. ^15,16^ These models quantify risk through a set of parameters — a combination of fixed (measurable risk factors) and random effects.^15^ Random effects defined with conditional autoregressive (CAR) priors help in risk estimation, integrating unmeasured confounding factors (underlying social contexts that are difficult to measure/unavailable) and their spatial dependency between areal units, as well as temporal dynamics. ^15^ This not only helps to reduce bias in risk estimation but is instrumental in integrating and exploring how heterogeneous social contexts shape patterns of risk over space and time. ^15^ The insights that random effects offer also hinge on its specification, such as the assumptions of how social context is spatially structured. ^15^

The current literature that apply Bayesian hierarchical model with CAR priors used to study COVID-19 and associated factors ^4,16–20^ predominantly use the Intrinsic CAR model proposed by Besag (1974) and the Besag, York, and Mollie (BYM) and their derivatives. ^21–24^ While beneficial in attempting to address spatial dependency, these models assume a single global autocorrelation across a region, overlooking subregional variations. ^25^ This misses the opportunity to integrate complex and localized social structures that might be connected to health outcomes, including COVID-19 risk.

To address gaps in the literature, this study uses spatio-temporal Localized Conditional Autoregressive Models (ST-LCAR) to assess the associations between socioeconomic and socio-demographic factors and the incidence of COVID-19 across the first four waves of the pandemic in Toronto. Piecewise intercepts highlight neighbourhood-level differences in COVID-19 risk linked to varying local social and temporal contexts.

## METHODS

### Study Location and Population

Toronto is Canada’s largest city with nearly 2.8 million residents.^26^ During the 2016 census, the City of Toronto was split into 96 Forward Sortation Areas (FSAs), which are geographical units designated by Canadian Postal codes. The FSA was chosen as the geographical unit of interest to account for testing and subsequent vaccination information collected at this areal unit. More details about the social demographic factors of the city are found in Table 1 and Figure 1

**Figure 1.**
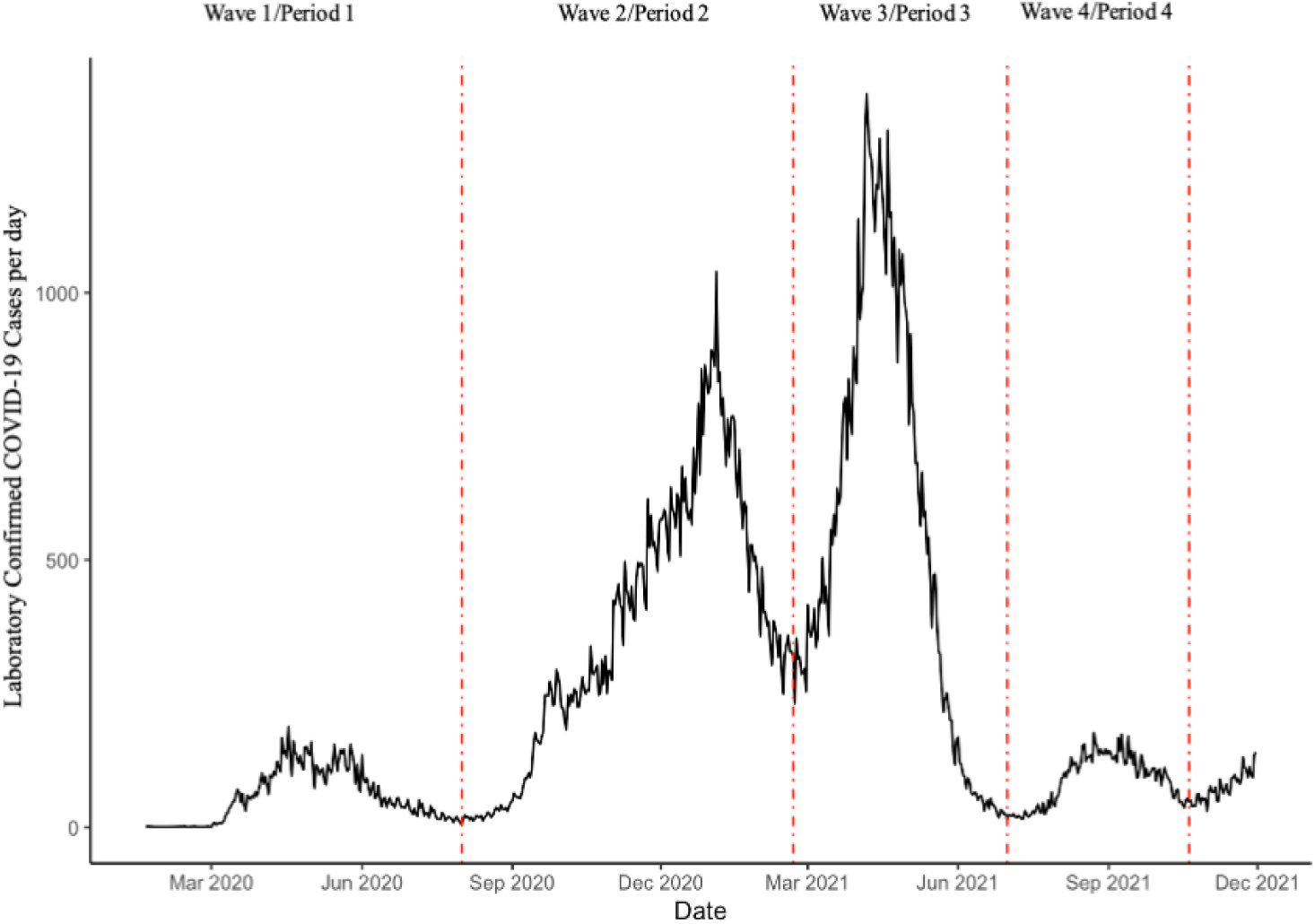
Daily Laboratory Confirmed COVID-19 cases in Toronto from January 2020-December 2021. Waves of COVID-19 are indicated based on the patterns of peaks and valleys in case counts

**Table 1.** Descriptive Statistics of the City of Toronto Using 2016 Canadian Census. ^19^

|  |  |
| --- | --- |
| **Total population in Toronto** | 2,794,356 |
| **Median Population per Forward Sortation Area** | 24,866 [IQR:18,531 – 36,933] |
| **Age** |  |
| Median Age in Toronto (years) | 40.5 [IQR: 39.3 – 42.4] |
| Proportion of Population over 65 years | 15% [IQR: 13% - 18%] |
| **Proportion of Residents that are Biologically Male** | 48% [IQR: 47% – 49%] |
| **Sociodemographic/Socioeconomic Characteristics** |  |
| Median after-tax income of households in 2015 (CAD) | 59,286 [IQR: 51,676 – 68,232] |
| Median percentage of population that identify as visible minorities | 45% [IQR: 29% – 66%] |
| Median percentage of population that identify as immigrants | 45% [IQR: 33% – 55%] |
| Median percentage of the population aged 25-64 with post-secondary education | 72% [IQR: 60% – 83%] |
| Average number of residents living in a household | 2.43 [IQR: 2.18 – 2.70] |
| **Median COVID-19 cases per week in each wave** |  |
| Wave 1 (January 20, 2020 – July 31, 2020) | 377 [IQR: 70.5 – 917] |
| Wave 2 (August 1, 2020 – February 19, 2021) | 2,314 [IQR: 834 -3657] |
| Wave 3 (February 20, 2021 – July 2, 2021) | 2,850 [IQR: 1011 – 5754] |
| Wave 4 (July 3, 2021 – October 30, 2021) | 606 [IQR: 536 – 4356] |

### Temporal Component

The study period was divided into four time periods (see Table 1) reflective of the first four complete waves of the pandemic in Toronto (see Figure 1), which were defined as distinct periods of sustained and substantial increase in disease transmission towards a peak and then a decline.^27^ These wave trends attempt to capture the unique experiences (virus variants, public health interventions, changes in testing access) that characterize the rise and fall of cases counted in each wave. ^12,28^ The study period ends on October 30 and excludes subsequent COVID-19 waves, as government-sponsored COVID-19 testing was limited to a select population by December 31st 2021, affecting the ability to include complete testing data for those waves.^29^

### Data

Laboratory-confirmed COVID-19 cases for each FSA were identified from Ontario’s Public Health Case and Contact Management system, a provincial surveillance database used to manage and report all laboratory-confirmed COVID-19 cases and their contacts. Cumulative number of cases was calculated for each FSA per wave. Disease risk was presented as the ratio of observed to expected cases (SIR: standardized incidence ratio) to account for potential population structure differences across FSA, given that age was a covariate of interest. To calculate expected case counts for each FSA, age-specific COVID-19 case counts in the Canadian population were identified from publicly available weekly COVID-19 case counts provided by Health Canada.^27^

Sociodemographic and socioeconomic factors of interest were selected from the 2016 Canadian Census. In addition, testing rate and second vaccination dose administered per FSA were accounted for as access to testing and vaccination were variable throughout the pandemic. Furthermore, they were identified to have an association with the number of cases recorded and COVID-19 risk, respectively. ^30,31^ Testing rates per FSA were identified through de-identified Ontario Laboratories Information System (OLIS).^32^ Second vaccination dose administered was collected through COVaxON, a provincial database that manages and reports COVID-19 vaccine administration across the province. Vaccination doses were linked to locations where vaccinations were provided not where recipients reside, given the nature of the deidentified COVaxON data. Vaccination was also logarithmically transformed as data was extremely left skewed.

### Model Selection

An initial set of socio-demographic and socioeconomic factors of interest was selected based on literature review of spatial studies in Toronto (see Table 1) ^1,2,4–6^ Given the potential for multicollinearity between closely related variables, a correlation plot was used to identify correlation between variables then followed with a Bayesian Poisson Regression model without any autoregressive terms to assess the variable inflation factor (VIF) for variables included in the model. VIF quantifies how strongly variables are correlated with others in the model. ^33^ Variables that were collinear or had a VIF greater than 5 (as a conservative choice) were removed ^33^ By utilizing both the correlation plots and variance of inflation factors, a finalized set of variables was selected to minimize collinearity. The Bayesian Poisson model with the finalized set of variables was performed to provide a baseline to compare how well the subsequent CAR models address spatial autocorrelation.

### Localized Spatial and Temporal Conditional Autoregressive Model (ST-LCAR)

The analysis follows methods previously outlined by Lee & Lawson (2016). ^25^

Risk (R_it_) of COVID-19 per FSA (*i*) and wave *t* were modelled with a Poisson distribution where Y_it_ is the observed number of COVID-19 cases in each FSA at wave *t*. The number of expected cases for each FSA (E_it_) was used as an offset for the Poisson regression.

The log relative risk of COVID-19 is given as

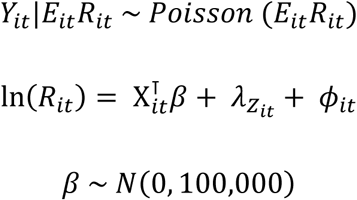

Where the log risk of COVID-19 over space and time 𝑅_𝑖𝑡_is a function of the covariates of interests Χ_𝑖𝑡_ with regression parameters 𝛽, and random effects, which includes spatio-temporal dependency 𝜙_𝑖𝑡_ and a piecewise intercept 𝜆_𝑍𝑖𝑡_. Adding the localized component to the model accounts for step changes in the spatial and temporal autocorrelation landscape that could result from localized social context not captured by the regression covariates.

The spatio-temporal random effect for each areal unit 𝜙_𝑖𝑡_ is modelled using a first-order conditional autoregressive (CAR) prior distribution that captures the globally smooth spatio-temporal autocorrelation in the data. Taken together, the vector of random effects (𝜙 = 𝜙_1_ …, 𝜙_𝑇_) across the City of Toronto over time is assumed to follow a Gaussian multivariate distribution.

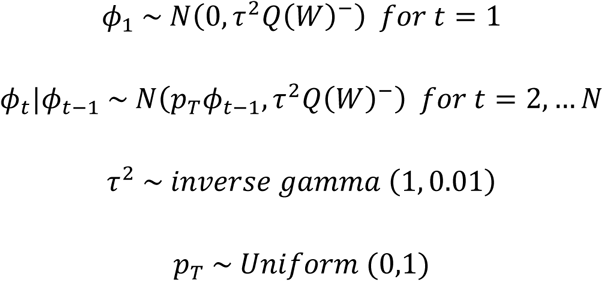

The temporal autocorrelation is induced via the mean with 𝑝_𝑇_ (temporal dependence parameter) ranging from 0 to 1 depending on the strength of temporal autocorrelation. Spatial autocorrelation is induced via the covariance matrix (𝜏^2^𝑄(𝑊)^−^) -- a combination of the variance of random effects (𝜏^2^) and the precision matrix (Q(W) which combines the neighborhood structure and the queen contiguity weighting matrix for neighborhoods). A spatial dependence parameter is usually included in the covariance matrix of other Bayesian Hierarchical CAR models; however, for this ST-LCAR model, this parameter is simplified to 1 to assume strong spatial dependence so that piecewise component (𝜆_𝑍𝑖𝑡_) can capture the localized clustering and step changes.

The piecewise 𝜆_𝑍𝑘𝑡_ clustering intercept

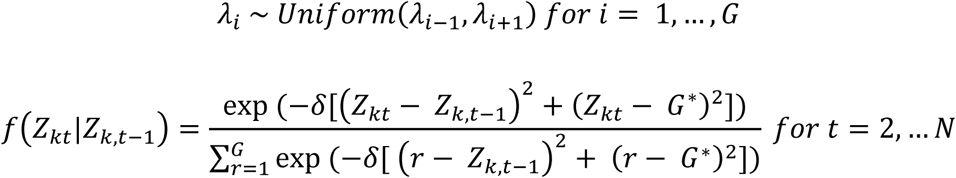

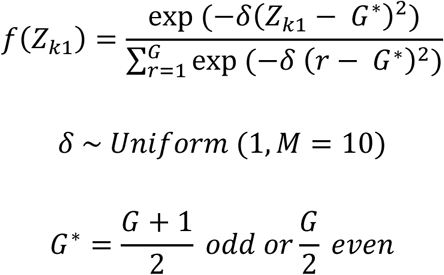

has at most *G* distinct levels 𝜆 = 𝜆_1_ … . 𝜆_𝐺_ where 𝑍_𝑖𝑡_ controls the assignment of each FSA at wave *t* to one of the *G* intercept levels. *G* is not necessarily the maximum number of distinct final intercepts, because the model can estimate fewer intercepts. This is because the prior distribution of 𝑍_𝑖𝑡_ is a Markov model with a penalizing term 𝛿(𝑍_𝑘_ − 𝐺^∗^)^2^ that includes the shrinking parameter 𝛿 so that FSAs are only in extremely high or low risk values if supported by the data.

For the analysis, an initial *G* value of 3 was selected to separate high-risk and low-risk FSAs from average risk. *G* Values were not estimated in the model as Lee & Lawson (2016)^25^ highlight computational constraints with this additional step. Weekly informative inverse gamma, uniform and Gaussian priors were specified for the regression parameters, precision parameter, and shrinking parameter.

### Sensitivity Analysis

To assess the robustness of their model, Lee and Lawson (2016)^25^ compared the results from their model to a popular ST-CAR model without the piecewise intercepts, proposed by Rushworth et. al., (2014)^24^. The ST-LCAR model was also built as an extension of the Rushworth et. al., (2014)^24^ model. This approach was also adopted for this paper. Specifically, the goal was to look for changes in estimated relative risks for each variable, model fit and compare how well spatial and temporal autocorrelation were addressed. CARBayesST ^24^ and rstanarm^25^ packages in R version 4.3.2 were used to carry out analysis.

### Spatial and Temporal Autocorrelation

A permutation test of Moran’s I statistics was used to assess whether models adequately addressed spatial autocorrelation at each time interval, while a Ljung Box^34^ test was used to assess whether the model addressed temporal autocorrelation across all areal units. P-values greater than 0.05 for both tests indicate that the model adequately addressed autocorrelation. ^26^ P-values were compared across models to gauge improvements in addressing residual spatial and temporal autocorrelation.

### Inferences, Model Fit and Convergence

All models were fitted with Markov chain Monte Carlo (MCMC) simulation with Gibbs sampling. Convergence of Markov Chains was identified through trace plots and convergence diagnostic proposed by Geweke (1992).^15^ Model fit was determined with the Deviance Information Criterion (DIC), the effective number of parameters (pD), and the Watanabe-Akaike Information Criterion.^15,35^ The best model minimizes the DIC, pD and the WAIC.

## RESULTS

### Model Selection

Most socioeconomic and sociodemographic factors displayed some level of collinearity, with visible minority and immigrant proportion being highly correlated (Pearson correlation coefficient > 0.75) See Figure 2. ^36^

**Figure 2.**
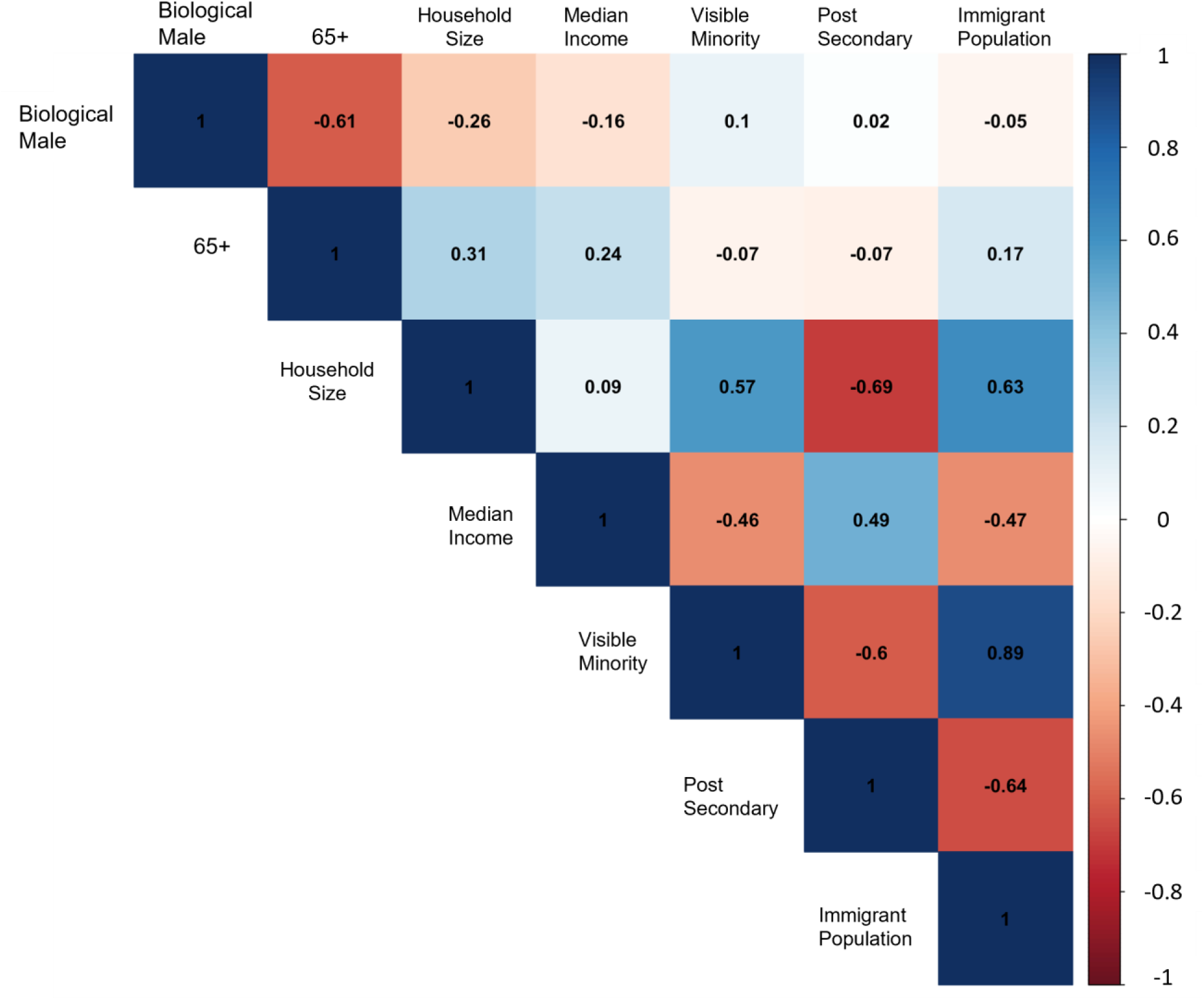
Correlation Plot depicting the collinear relationships between initial sociodemographic and socioeconomic variables of interest.

The immigrant variable was removed while visible minority variable was maintained with the intent of capturing individuals who self-identified as visible minorities but did not identify as immigrants. A Bayesian Poisson Regression model without any autoregressive terms was also generated to calculate variance of inflation (VIF) factor to assess multicollinearity for the remaining covariates when included into the model. ^33^ The variable concerning the average number of residents per household had a VIF larger than 5 and was thus removed. ^33^ Refer to Figure 3 for a spatial representation of the final set of variables.

**Figure 3.**
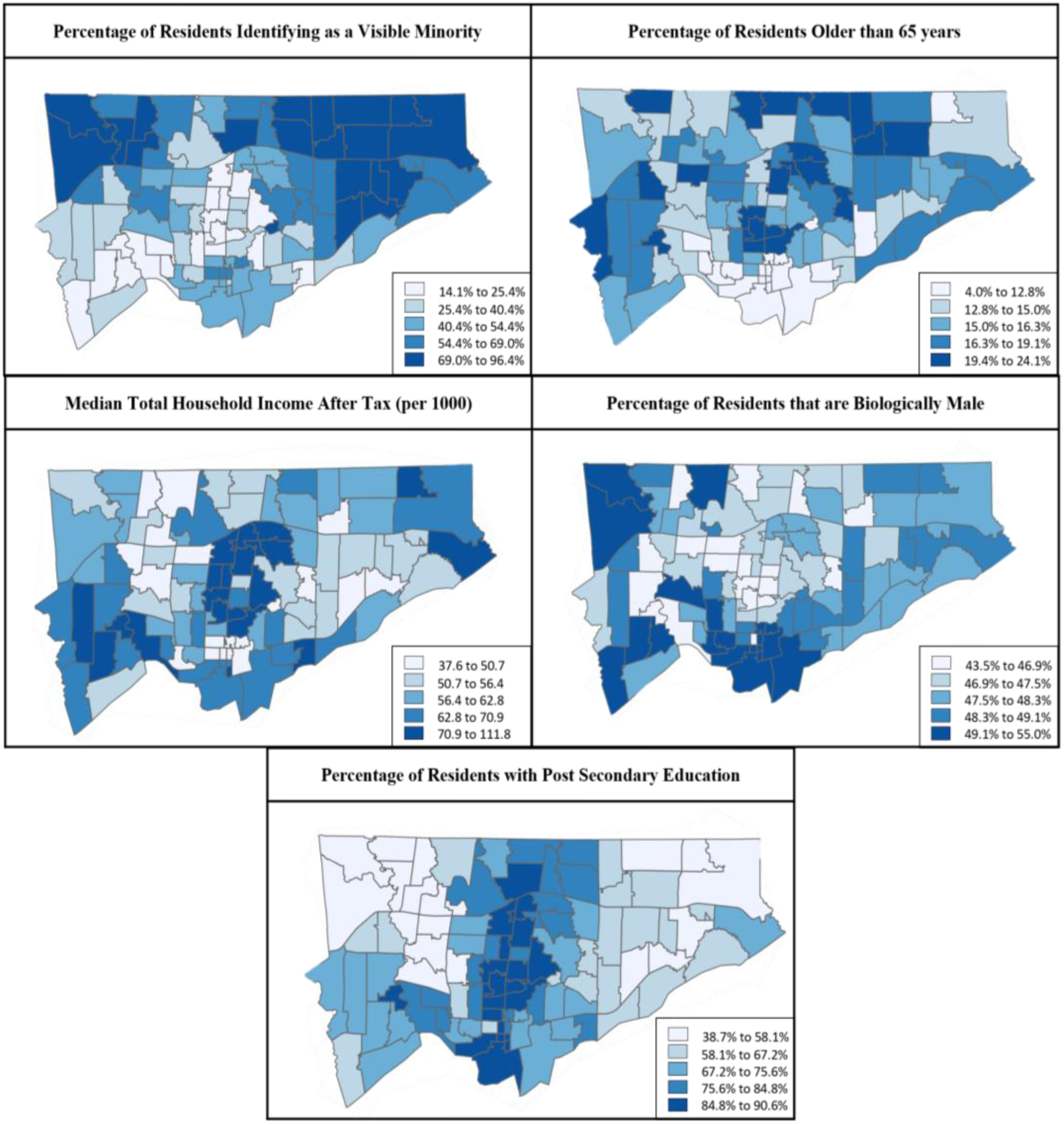
Spatial distribution of finalized sociodemographic and socioeconomic factors across the City of Toronto based on 2016 Canadian census

### COVID-19 Relative, Sociodemographic and Socioeconomic factors

The localized ST-LCAR model helps quantify the association between social context and COVID-19 relative risk through enumerated sociodemographic and socioeconomic factors and a and localized components (piecewise intercepts) that integrates unmeasured local social context/potential step changes in COVID-19 relative risk over space and time.

A 10-percentage point increase in testing rate (RR 1.16 (1.13 - 1.20)) was associated with increased COVID-19 relative risk. A 10-percentage point increase in the proportion of visible minorities in an FSA was associated with a moderately increased COVID-19 relative risk (RR 1.03 (1.02 – 1.07)). A 10-percentage point increase in the proportion of residents in an FSA who were older than 65 or identified as biological male were also associated with higher relative risk point estimates, though credible intervals did include 1. On the flip side, a 10-percentage-point increase in the proportion of residents with post-secondary education in an FSA (RR 0.78 (0.75 – 0.82)) was associated with a decreased COVID-19 relative risk. Income, and vaccination were not associated with COVID-19 relative risk.

FSAs are assigned to different groups/intercepts depending on their inherent COVID-19 relative risk over space and time. The localized model identified three step changes/relative risk groups (RR1: 0.50, RR2: 1.11, RR3: 2.14) (see Figure 4). From wave 1 onward, higher relative risk clusters FSAs (RR3) do change throughout waves, depicting a changing landscape of inherent COVID-19 risk values potentially highlighting the connection between social context and temporal changes.

**Figure 4.**
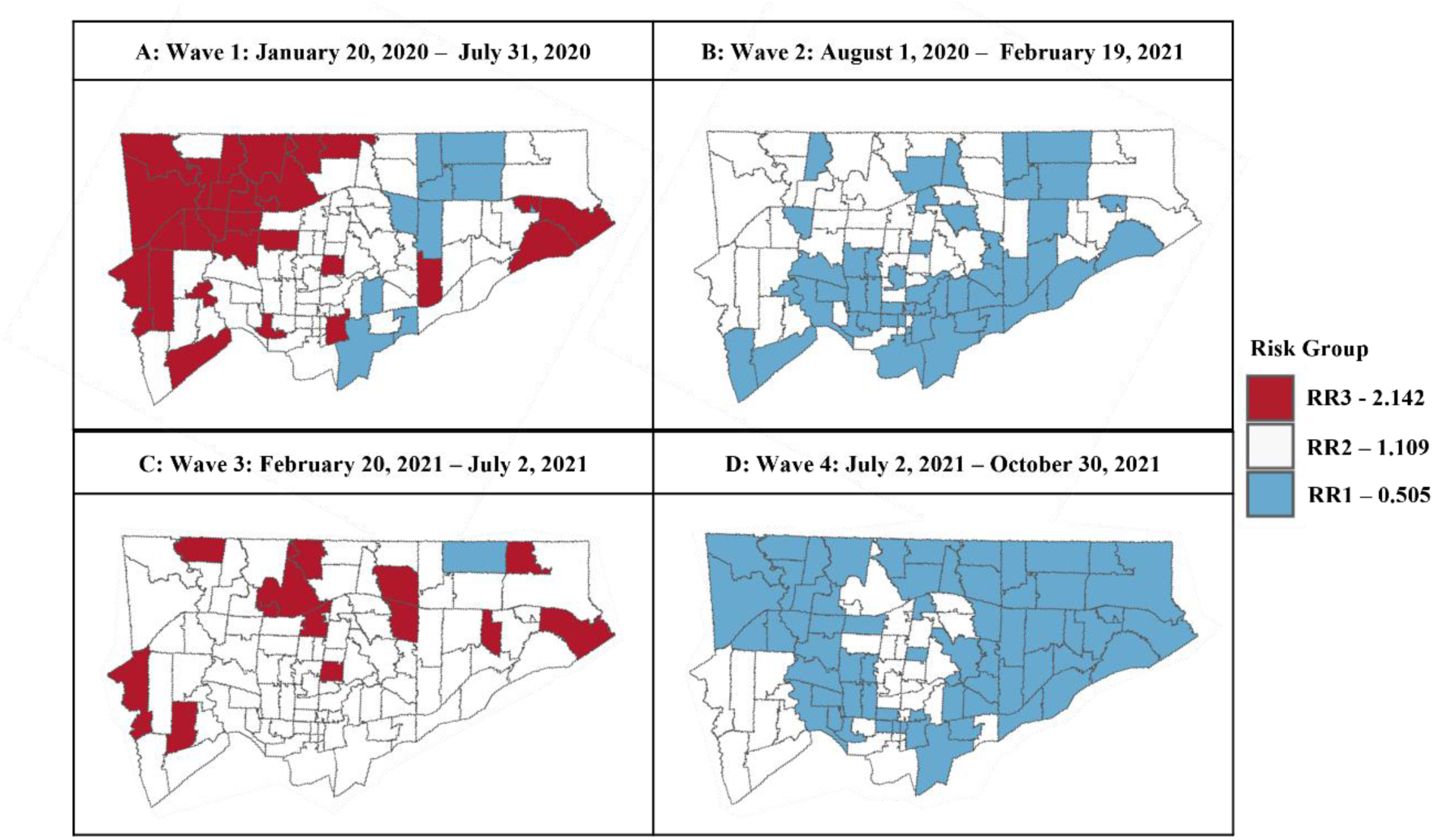
Relative Risk Groups identified from ST-LCAR Model. RR3 groups FSAs with the highest relative risk of COVID-19 across space and time while RR1 groups FSA with the lowest relative risk group across space and time.

### COVID-19 Relative Risk in Toronto

Relative risks of COVID-19 across FSAs during each wave and cumulatively across all waves estimated by the ST-LCAR model combining both fixed and random effects are displayed in Figure 5. Overall, as seen in Figure 5A, FSAs in the northwestern corner of Toronto had higher relative risk of COVID-19, with some FSAs having 3-time higher relative risks than the rest of the FSAs in Toronto throughout the four waves. The lower relative risk of COVID-19 was concentrated in the center of the city. When one takes a closer look through each wave (Figure 5B - 5E), there is more nuance across waves. While relative risks were high in the northwestern corners of the city, overall relative risk decreased over time. The patterns shift by the fourth wave. As compared to the rest of the waves, the relative risk for COVID-19 in the fourth wave is lower with slightly elevated risk in FSAs across the western border of the city.

**Figure 5.**
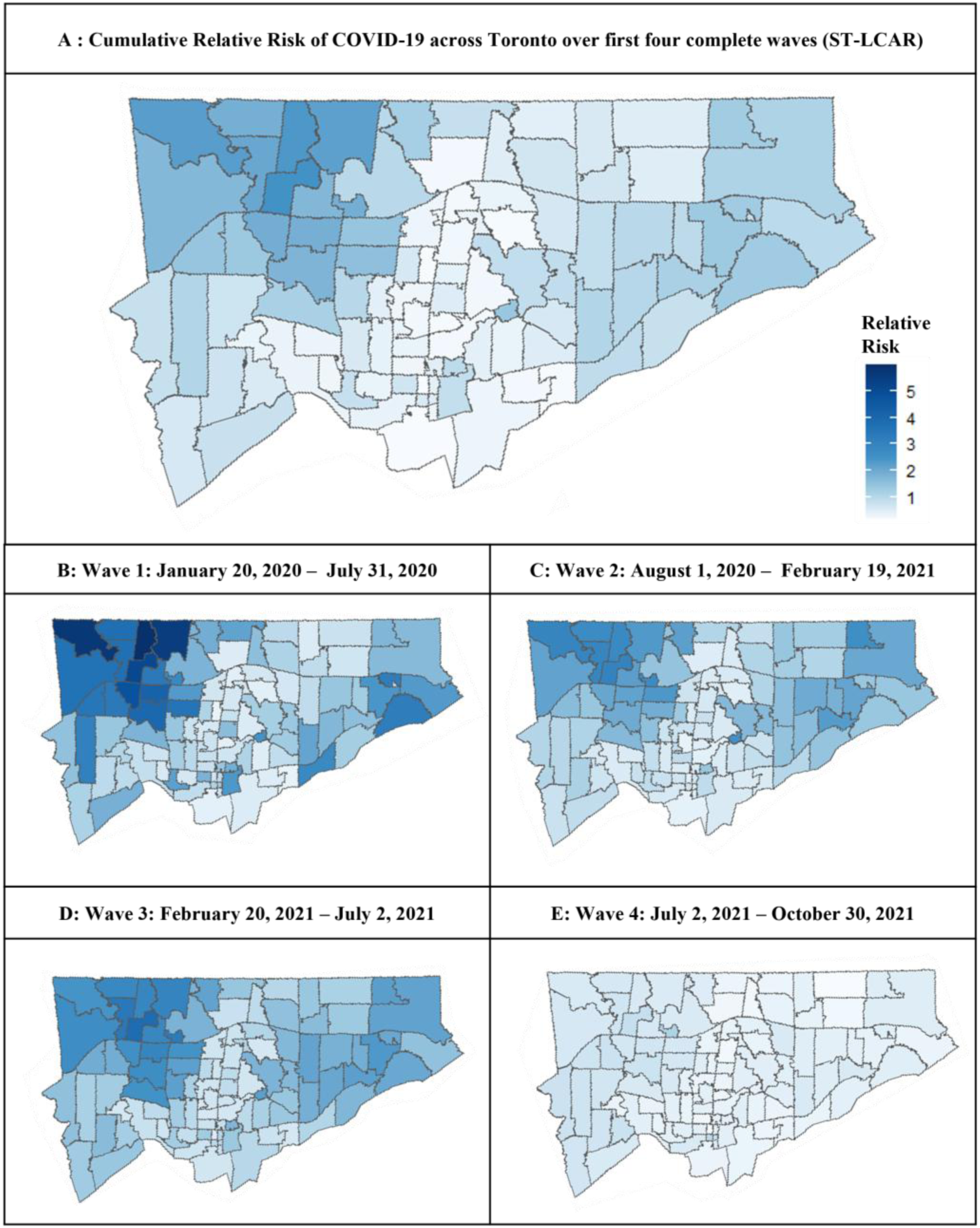
Relative Risk of COVID-19 in Toronto Across the first four complete waves from January 2020 to October 2021

When the results from the ST-LCAR produced maps (Figure 5) are overlaid with the distribution of sociodemographic and socioeconomic factors displayed in Figure 3, potential connections arise. Namely, a potential increased relative risk in FSAs with a higher concentration of visible minorities and a potential decreased relative risk in FSAs with a higher percentage of post-secondary educated residents. However, there is variability in potential associations.

In parallel, comparing the relative risk maps generated from full models (Figure 5) with the maps (Figure 4) depicting inherent, localized spatial-temporal risk can offer further insights into how well our enumerated sociodemographic and socioeconomic factors capture the connection between social context and risk. Unlike waves 3 and 4, the relative risk patterns in the first 2 waves (5A and B) mirror those in Figure 4, suggesting that the enumerated sociodemographic and socioeconomic factors may not fully explain the relative risk landscape in these time periods.

### Sensitivity Analysis

To ensure the robustness of the localized model, results generated were compared with an ST-CAR model alternative ST-CAR model described by Rushworth et al. 2014 ^24^ without a piecewise intercept (Table 2). Sensitivity analysis yielded similar trends, but with greater uncertainty for some covariates. Furthermore, Rushworth et al. (2014)^24^ The model does have a larger DIC and pD values as compared to the primary localized model, which might indicate moderately less fit. Relative risk maps generated from the sensitivity analysis are shown in Supplemental Figure 1; they resemble those from the ST-LCAR model in Figure 5.

**Table 2.** Covariate Estimate from Primary Analysis model and Sensitivity Analysis using Spatio-Temporal (Localized) Conditional Autoregressive Models.

| Variables | Bayesian Poisson Model with no Random Effects | Primary Model – ST-LCAR Model<sup>25</sup> | Sensitivity Model – ST-CAR Rushworth Model<sup>24</sup> |
| --- | --- | --- | --- |
| **Testing and Vaccination** |  |  |  |
| Per 10-pp<sup>1</sup> increase in Testing Rate per FSA | 1.11 (1.10 – 1.11) | **1.16 (1.13 - 1.20)** | 1.13 (1.09 – 1.18) |
| Per 10-pp<sup>1</sup> increase in vaccine doses administered per FSA | 0.99 (0.99-0.99) | **0.99 (0.99 – 0.99)** | 0.99 (0.99 – 0.99) |
| **Biological Factors** |  |  |  |
| Per 10-pp<sup>1</sup> increase the population of resident older than 65 | 1.00 (0.98 – 1.02) | **1.10 (0.97 - 1.26)** | 1.00 (0.83 – 1.21) |
| Per 10-pp<sup>1</sup> increase in the population of residents biologically male | 0.82 (0.79 – 0.86) | **1.26 (0.92 – 1.72)** | 1.10 (0.73 – 1.65) |
| **Sociodemographic/Socioeconomic Characteristics** |  |  |  |
| Per 1000 CDN increase in median after-tax income of households in 2015 | 0.99 (0.99 – 0.99) | **0.99 (0.99 – 0.99)** | 0.99 (0.99 – 1.00) |
| Per 10-pp<sup>1</sup> increase in the population of visible minority resident | 1.03 (1.03 – 1.03) | **1.03 (1.02 – 1.07)** | 1.01 (0.98 – 1.04) |
| Per 10-pp<sup>1</sup> increase in the population of residents with post secondary education | 0.77 (0.77 – 0.78) | **0.78 (0.75 – 0.82)** | 0.77 (0.73 – 0.82) |
| Relative Risk Group 1 |  | **0.51 (0.49 – 0.53)** |  |
| Relative Risk Group 2 |  | **1.11 (1.08 – 1.14)** |  |
| Relative Risk Group 3 |  | **2.14 (2.05 – 2.24)** |  |
| **Model Fit** |  |  |  |
| DIC<sup>15</sup> |  | **3292** | 3534 |
| pD<sup>15</sup> |  | **106** | 376 |
| WAIC<sup>35</sup> | 36223 | **3491** | 3428 |
| Moran's I pValue | 9.99 x 10<sup>-5</sup> | **0.99** | 0.87 |
| Median Ljung Box<sup>34</sup> pValue per FSA | 0.33 | **0.40** | 0.37 |

<sup>1</sup> 10-pp: 10-percentage point increase

## DISCUSSION

A spatio-temporal localized conditional autoregressive model was used to assess the relationship between COVID-19 risks and socioeconomic and sociodemographic factors, considering the underlying localized social context on COVID-19 relative risk and temporal dynamics. The localized model identified an increased relative risk of COVID-19 in the northwestern corner of the city, highlighting changes in the relative risk landscape during the fourth wave. Focusing on the potential relationships between sociodemographic and socioeconomic factors and COVID-19 disease risk, population differences in biological sex, age, education level and self-identified visible minority status per FSA were associated with differences in estimated COVID-19 disease risk, although the strength of these relationships varied. Only visible minority status and education showed consistent directions of associations. Furthermore, the ST-LCAR model provided a visual representation of the localized risk structure through the piecewise intercept connected to underlining differences in local social contexts and temporal dependency. This helps to identify locales with higher-than-average COVID-19 relative risk or lower-than-average COVID-19 risk, the step changes in these relative risk patterns over space, and how this pattern also changes over time.

The results from the localized model have been corroborated by the current literature on COVID-19 in Toronto. Other studies have identified an increased risk of COVID-19 in the northwestern corners of the city, with clustering more pronounced in areas with a larger proportion of visible minorities and lower educational attainment. ^1–4^ Research denotes that visible minorities are likely to live in low-income, densely populated housing, and work low-income and/or “essential” jobs with fewer opportunities to work from home or social distance and increased risk of exposure. ^1,3,10,37,38^ Education levels also affect job opportunities for individuals, with a post-secondary education positioning individuals to access jobs with more flexibility and the ability to work from home. ^1,10^

These enumerated patterns are just pieces of a very complex puzzle of interacting social contexts that shape health inequalities. Social context in the city differs along ethno-cultural, economic and environmental conditions. ^8,10^ These differences can be seen in neighbourhood conditions, housing access and quality, work opportunities, social interactions and access to resources, all of which are associated with COVID-19 risk in many ways, might may be difficult to fully enumerate. ^39^ Neighbourhoods that are structurally disadvantaged often lack access to health-promoting resources such as affordable healthy food, clean air, and a safe walkable environment, which are connected to COVID-19 risk and adverse outcomes through comorbidities.^10,40^ They are densely populated with lower-income, racialized residents living in poorly maintained high-rise apartments with poor air quality, which, in connection to a multigenerational living environment, can collectively heighten transmission, either through increased contact or lingering aerosols shed by infected individuals. ^9,10,41^ These populations rely on public transit as the primary mode of transportation, further increasing contact with others.^1^ Public health interventions recommended early in the pandemic (social distancing, isolation, and working from home) would have been challenging to adhere to in these environments.^13^

Additionally, the difference in ethno-cultural composition across neighbourhoods in the city may have further shaped COVID-19 patterns. There were neighbourhoods in the city, both with high proportion of visible minority and immigrant residents, yet with different relative risk patterns.

Toronto’s diverse landscape has been described to be segregated along race, creating ethnic enclaves each rooted in distinct histories, culture and social cohesion. ^8^ It has been identified that Black Torontonians were more than 7 times likely than East Asian counterparts to contract COVID-19, with higher seropositivity to SARS-CoV-2 infection in neighbourhoods with a high proportion of Black residents. ^39^ Black residents also had a lower rate of COVID-19 vaccine uptake. ^42^ Different racial experiences connect to institutional and structural racism, cultural context and social positioning, and these patterns were potentially visible in the spatial patterns of COVID-19 relative risk ^43,44^ Unfortunately, a lack of comprehensive race-based COVID-19 data has made it challenging to further investigate differential COVID-19 experiences.^45^

As COVID-19 remains endemic in Toronto, interventions to reduce transmission and improve pandemic preparedness must recognize the complex ways in which differential social contexts shape COVID-19 susceptibility and exposure. Interventions could include updating housing policies that consider air quality, developing culturally informed public health messaging around COVID-19 preventative measures and seasonal COVID-19 vaccination, strengthening collaboration with community health organizations, creating policies in pandemic preparedness plans that formalize emergency paid sick leave with accompanying financial supports to stay home when sick, and integrating an equity focused lens in public health surveillance by expanding sociodemographic data collection with COVID-19 health outcomes.^10,46,47^

### Limitations

The benefit of using spatio-temporal localized Bayesian hierarchical models is recognizing the presence of unmeasured, and complex underlying local social systems that may be associated with COVID-19 relative risk. However, this strength is also a limitation, as while it is possible to identify a potential connection between unmeasured confounding factors and the relative risk estimate, one can only hypothesize about what these underlying contexts might be, especially without access to rich data that aptly describes the differing social contexts in the city.

Recognizing that COVID-19 testing access, provincial testing capacity, and reporting may have shaped testing uptake, laboratory-confirmed COVID-19 cases may not capture the true patterns of infections. ^12,48^ The influence of variable testing uptake on COVID-19 case counts was accounted for by adjusting all models for testing rates and including a temporal component in this study. Because testing affects case detection, a follow-up study will investigate testing patterns and the factors that shaped testing rates.

With any spatial study, there is a concern about modifiable areal unit problem (MAUP). The ability to address MAUP was limited, as testing and vaccination data were available only at the FSA level. Unlike other levels of census area demarcation, the FSA’s structure is not defined by the socio-demographic characteristics of residents and can thus have a mix of neighbourhoods that differ along cultural, economic and physical characteristics. A heterogeneous FSA may underestimate associations between COVID-19 risk and sociodemographic or socioeconomic factors. Furthermore, marginalization indices that have been created to integrate numerous census-based data to provide more detailed social context beyond individual sociodemographic and socioeconomic factors were also not available at the FSA level.^49^ Given the possibility that variable testing could directly affect how many COVID-19 cases were accounted for and the necessity to consider vaccination in describing the COVID-19 experience, it was more important to favor the areal units that offered the ability to adjust for testing and vaccination.

While the localized model provided useful insights into the relationship between socioeconomic and sociodemographic factors and disease risk, considering clustering introduced some pitfalls. The convergence of MCMC chains to estimate the piecewise intercept values (lambdas) was challenging (specifically trying to find the number of MCMC samples that allowed convergence) and required more computational power. This issue was not encountered with the sensitivity analysis, where all MCMC chains converged without much difficulty. There is a possibility that the additional parameters the localized model needed to estimate – spatial and temporal effects, piecewise intercepts, and hyperparameters that estimate these intercepts and effects- affected the efficiency of the model. The authors of the ST-LCAR model have recommended an alternative method to capture the localized difference in the weighting matrix -- an approach that might require more technical skills than the user-friendly ST-LCAR, and can be explored in subsequent research projects.^50^ The ST-LCAR model still instills confidence, as the results were corroborated in the literature.

## Conclusion

This study demonstrates that differential spatio-temporal patterns of COVID-19 across the City of Toronto cannot be understood without considering the pandemic’s temporal dynamics alongside embedded structural inequities that shape the city’s diverse ethno-cultural, environmental, and economic contexts. These different contexts are complex and define the neighbourhood, housing, working environment and resources that may be associated with infectious disease transmission and susceptibility. Structurally disadvantaged neighbourhoods were associated with increased COVID-19 relative risk, with sociodemographic and socioeconomic factors such as visible minority status and educational attainment associated with this result. As COVID-19 remains endemic, these findings underscore the need for public health interventions and pandemic preparedness that integrate an equity-focused lens, developing public health strategies that consider the diverse local social contexts that shaped COVID-19 experiences across the city.

## Author’s contributions

AA, MC, EG, ART and DNF contributed to the conceptualization, design of the study, drafted and/or substantially revised the work, have approved the submitted version and take responsibility for the submitted work. AA analyzed and interpretated the spatial and temporal data. DNF and AT acquired necessary data for the study.

## Ethics Approval

This study has been approved by the University of Toronto Health Sciences Research Ethics Board (protocol number 44167, approved March 17, 2023). This study conducts a secondary analysis of public health surveillance data that is deidentified and collected under Ontario’s Health Protection and Promotion Act. The ethics committee waived the need for informed consent.

## Data Availability

COVID-19 case counts for the City of Toronto are publicly available through the City of Toronto Open Data Portal. Versions of COVID-19 testing and vaccination data are publicly available through the Government of Ontario Data Catalogue. Census Data is publicly available through Statistics Canada.

https://open.toronto.ca/dataset/covid-19-cases-in-toronto/

## SUPPLEMENTAL INFORMATION

**Supplemental Figure 1.**
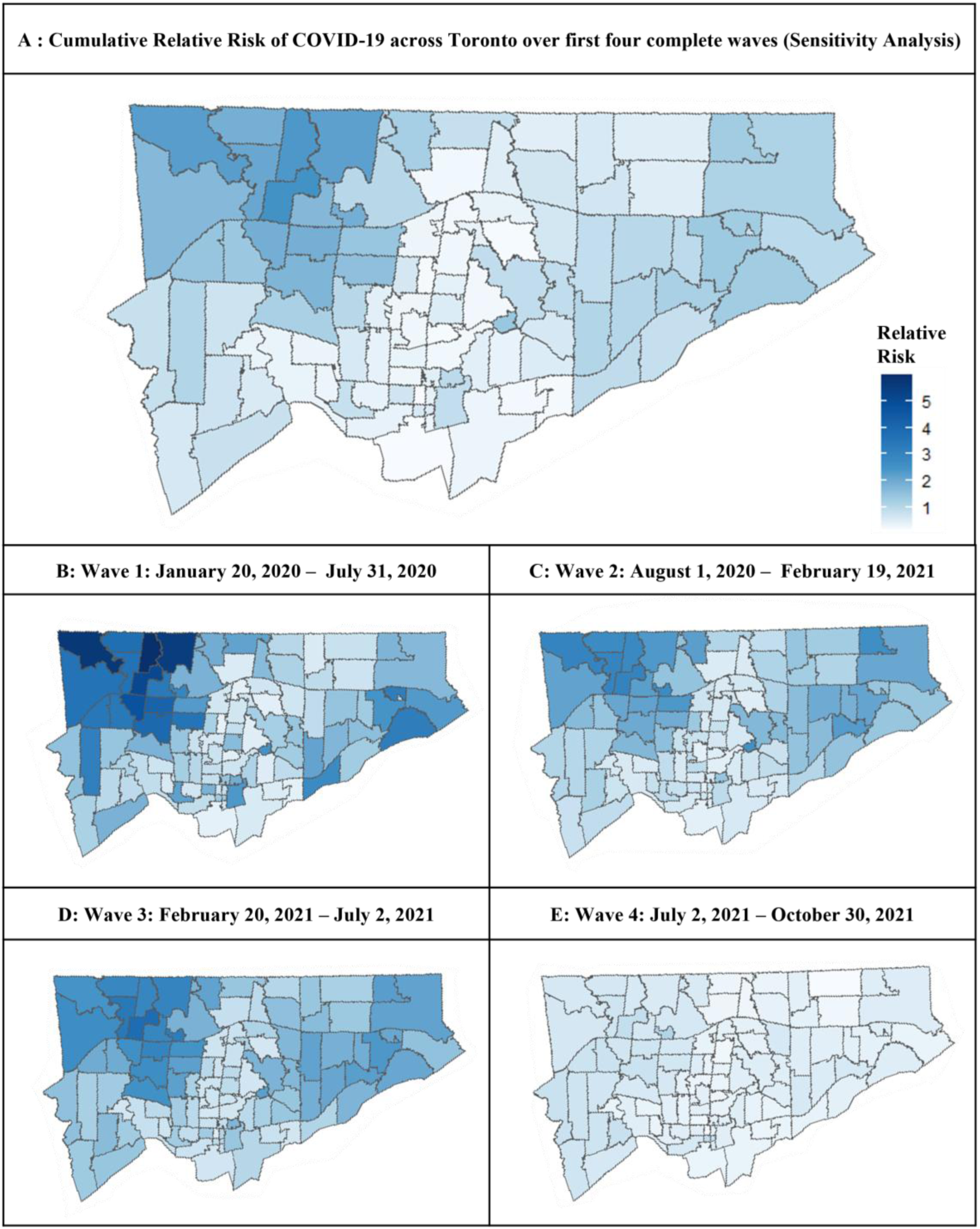
Relative Risk of COVID-19 in Toronto across the first four complete waves from January 2020 to October 2021 calculated for sensitivity analysis using a ST-CAR model without a piecewise intercept.

